# Modelling the potential impact of social distancing on the COVID-19 epidemic in South Africa

**DOI:** 10.1101/2020.04.21.20074492

**Authors:** F. Nyabadza, F. Chirove, W. Chukwu, M.V. Visaya

## Abstract

The novel coronavirus (COVID-19) pandemic continues to be a global health problem whose impact has been significantly felt in South Africa. Social distancing has been touted as the best form of response in managing a rapid increase in the number of infected cases. In this paper, we present a deterministic model to model the impact of social distancing on the transmission dynamics of COVID-19 in South Africa. The model is fitted to the currently available data on the cumulative number of infected cases and a scenario analysis on different levels of social distancing are presented. The results show a continued rise in the number of cases in the lock down period with the current levels of social distancing albeit at a lower rate. The model shows that the number of cases will rise to above 4000 cases by the end of the lockdown. The model also looks at the impact of relaxing the social distancing measures after the initial announcement of the lock down measures. A relaxation of the social distancing by 2% can result in a 23% rise in the number of cumulative cases while on the other hand increasing the levels of social distancing by 2% would reduce the number of cumulative cases by about 18%. These results have implications on the management and policy direction in the early phases of the epidemic.

## 1 Introduction

COVID-19 is an emerging respiratory infection, that is, it is a disease caused by a virus SARS COV-2 that has not been observed previously within a population or geographic location [1]. It spreads mainly via respiratory droplets produced when an infected person coughs or sneezes. The droplets are transferred to another individual through close personal contact such as touching or shaking hands, touching an object or surface with the virus on it and subsequently touching one’s face with contaminated hands [2]. People can also get the infection through breathing in droplets coughed by someone affected [3]. In South Africa, the emergence of COVID-19 was initially affected by human factors which included, international travel to infected countries and consequently local travels, human behaviour and crowding in cities, among others [5]. There are various ways that can be used to address, prevent, and combat the spread of COVID-19. These include regular and thorough cleaning of hands with alcohol based sanitizers, social distancing, avoiding touching one’s face, practising good respiratory hygiene, self isolation, quarantining, restricting travel, and lock-down [3]. There is currently no antiviral treatment or vaccine for COVID-19 and thus, most procedures are just supportive treatment such as provision of oxygen [5].

The first COVID-19 case in South Africa was identified on the fifth of March 2020 as an imported case from one of the hot-spots countries, Italy, in Europe. The number of imported cases continued to rise in the country until the 17^*th*^ of March when the first locally transmitted cases were identified. On this day the total number of cases recorded was 85 inclusive of eight (8) new locally transmitted cases [5]. The new cases came two days after the pronouncement of the national state of disaster by the president of the nation. On 26 March the nation enforced a 21-day national lockdown to restrict movement of persons during this period for which the regulation was in force and effect. The restriction largely enforced social distancing a measure to delay and prevent new locally and international infections [6]. Social distancing, means keeping space between yourself and other people outside of your home. This entails limiting close contact with individuals outside the household, in indoor and outdoor spaces [4]. If used effectively, it can delay the time to epidemic peak and reduce the overall number of cases, the number of cases at epidemic peak, and the total number of severe cases and deaths. The reduction of number of cases during the epidemic peak and the subsequent spread of cases over a longer time-period reduces the burden of health care systems thereby promoting effective case management and treatment [7]. Based on statistics, there are about 3 000 (out of about 7 000) critical care beds available between the public and private healthcare sectors reserved for critical COVID-19 patients [13]. A study done in 2017, showed that nationally, there were 1 hospital, 187 hospital beds and 42 surgical beds per 100 000 population [12]. South Africa has the most number of COVID-19 infections in the whole of Africa, with a total of 2272 cases as of April 13. Receiving over 21 million people annually with the capacity to process up to 28 million, O.R. Tambo International Airport in Johannesburg is Africa’s busiest airport. It is not surprising that South Africa’s first 77 cases were due to imported cases, with first 8 local transmissions reported on March 17, 2020. The South African government however acted fast and more decisively than most governments around the world–restricting travel from high risk countries just 9 days after detection of the first locally transmission case. Table 1 gives some of the important dates from the first case of COVID-19 in the country.

**Table 1:** COVID-19 Timeline in South Africa. The dates are for 2020.

| Date | Restriction/event |
| --- | --- |
| March 5 | first imported case recorded. |
| March 15 | President’s first speech addressing COVID-19. |
| March 16 | 72 ports of entry shut down. |
| March 17 | first local transmission. |
| March 18 | schools closed. |
|  | travel ban on foreign nationals from high-risk countries. |
| March 23 | announcement of national total lockdown. |
| March 26 | 21 day national lockdown begins. |
|  | movement between provinces, metros and district areas banned. |
|  | public transport for essential trips restricted to 05:00 to 09:00 and 16:00 to 20:00. |
| March 28 | first death recorded. |
| March 29, April 2 | first cases of infection in townships (Khayelitsha in the Western Cape and Alexandra in Gauteng, respectively) recorded. |
| April 1 | taxi industry regulations revised, passenger loading capacity, from 50% to 70% on minibus taxis. |
| April 2 | spaza shops and street vendors given green light to operate. |
| April 8 | extension of a further 14 day lockdown to April 30 announced. |

**Table 2:**
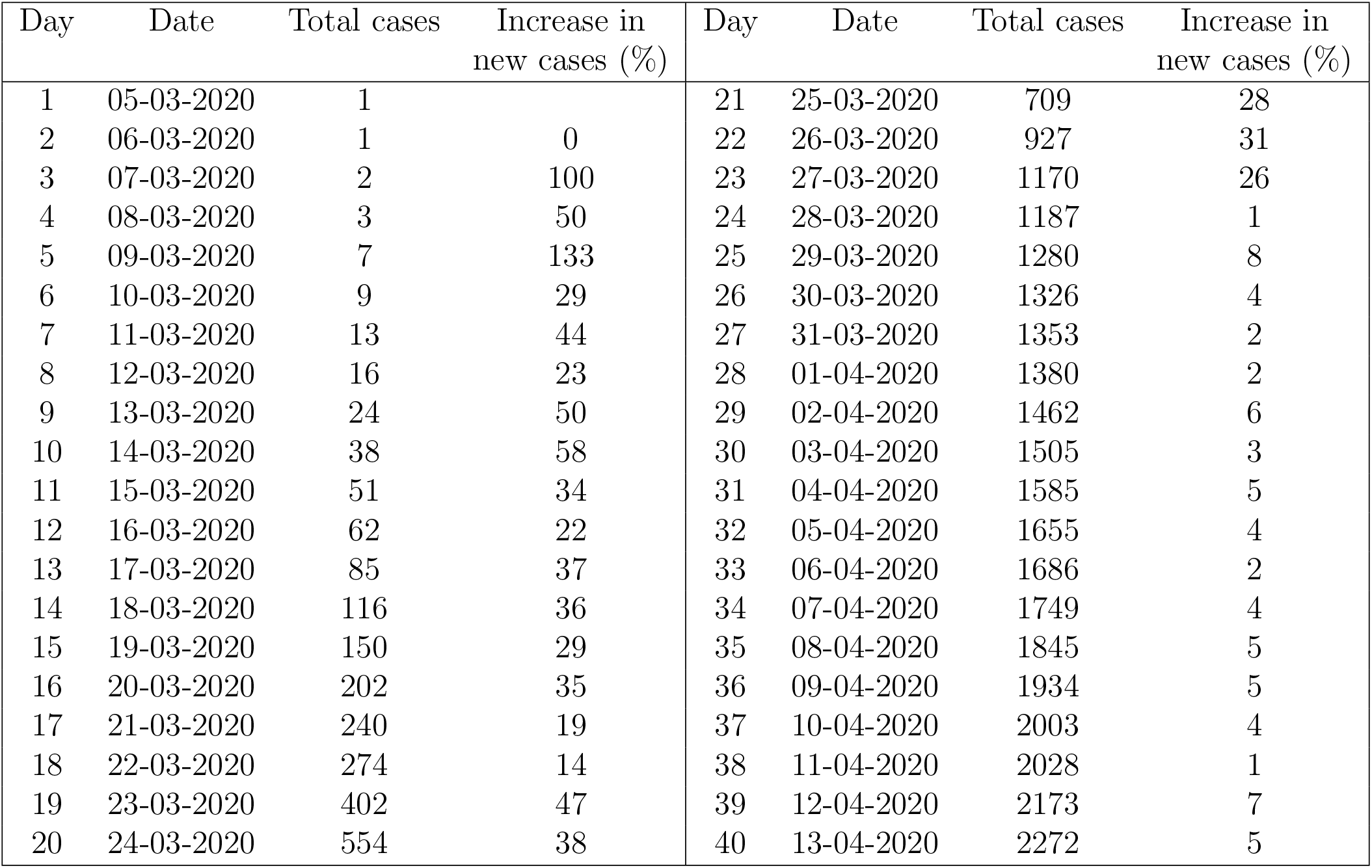
COVID-19 South Africa data [10, 11].

| Day | Date | Total cases | Increase in<br>new cases (%) | Day | Date | Total cases | Increase in<br>new cases (%) |
| --- | --- | --- | --- | --- | --- | --- | --- |
| 1 | 05-03-2020 | 1 |  | 21 | 25-03-2020 | 709 | 28 |
| 2 | 06-03-2020 | 1 | 0 | 22 | 26-03-2020 | 927 | 31 |
| 3 | 07-03-2020 | 2 | 100 | 23 | 27-03-2020 | 1170 | 26 |
| 4 | 08-03-2020 | 3 | 50 | 24 | 28-03-2020 | 1187 | 1 |
| 5 | 09-03-2020 | 7 | 133 | 25 | 29-03-2020 | 1280 | 8 |
| 6 | 10-03-2020 | 9 | 29 | 26 | 30-03-2020 | 1326 | 4 |
| 7 | 11-03-2020 | 13 | 44 | 27 | 31-03-2020 | 1353 | 2 |
| 8 | 12-03-2020 | 16 | 23 | 28 | 01-04-2020 | 1380 | 2 |
| 9 | 13-03-2020 | 24 | 50 | 29 | 02-04-2020 | 1462 | 6 |
| 10 | 14-03-2020 | 38 | 58 | 30 | 03-04-2020 | 1505 | 3 |
| 11 | 15-03-2020 | 51 | 34 | 31 | 04-04-2020 | 1585 | 5 |
| 12 | 16-03-2020 | 62 | 22 | 32 | 05-04-2020 | 1655 | 4 |
| 13 | 17-03-2020 | 85 | 37 | 33 | 06-04-2020 | 1686 | 2 |
| 14 | 18-03-2020 | 116 | 36 | 34 | 07-04-2020 | 1749 | 4 |
| 15 | 19-03-2020 | 150 | 29 | 35 | 08-04-2020 | 1845 | 5 |
| 16 | 20-03-2020 | 202 | 35 | 36 | 09-04-2020 | 1934 | 5 |
| 17 | 21-03-2020 | 240 | 19 | 37 | 10-04-2020 | 2003 | 4 |
| 18 | 22-03-2020 | 274 | 14 | 38 | 11-04-2020 | 2028 | 1 |
| 19 | 23-03-2020 | 402 | 47 | 39 | 12-04-2020 | 2173 | 7 |
| 20 | 24-03-2020 | 554 | 38 | 40 | 13-04-2020 | 2272 | 5 |

## 2 Methodology

### 2.1 Social distancing and the epidemic data

The *SEIR* model considered in this study considers the effect of social distancing so we are concerned with the mobility of South Africa soon after the lockdown. To get a sense of the general pattern of movement of the South African community before and after the lockdown, we use information from Google mobility reports [9]. Figure 1 depicts how visits and length of stay at different places change compared to a baseline (i.e the median value, for the corresponding day of the week, during the 5-week period Jan 3–Feb 6, 2020). Insights in these reports are created with aggregated, anonymised sets of data from users who have turned on the “location history” setting. The most recent report at the time of writing is April 11, and represents data about two to three days in retrospect, the time it takes to produce the reports. Categories that are useful to social distancing efforts as well as access to essential services are included in the report. It is evident from Figure 1 that the overall trend in South Africa has decreased after the lock-down (Mar 26). In particular, the behaviour in retail and recreation (-75%) and grocery and pharmacy (-46%) are similar, particularly a spike just before the lock-down as most people stocked up and did panic-buying. In both categories, there are still movement due to essential workers that need to go to work, with a decrease in activity during weekends. On the other hand, the opposite (complementing) trend between workplaces (-56%) and residential (+22%) can also be observed. With regards to the effect of social distancing to the COVID-19 cases, we can also observe from Figure 2 that the number of new infected cases have dramatically decreased after the lockdown, as depicted by the graph of the 7-day moving average.

**Figure 1:**
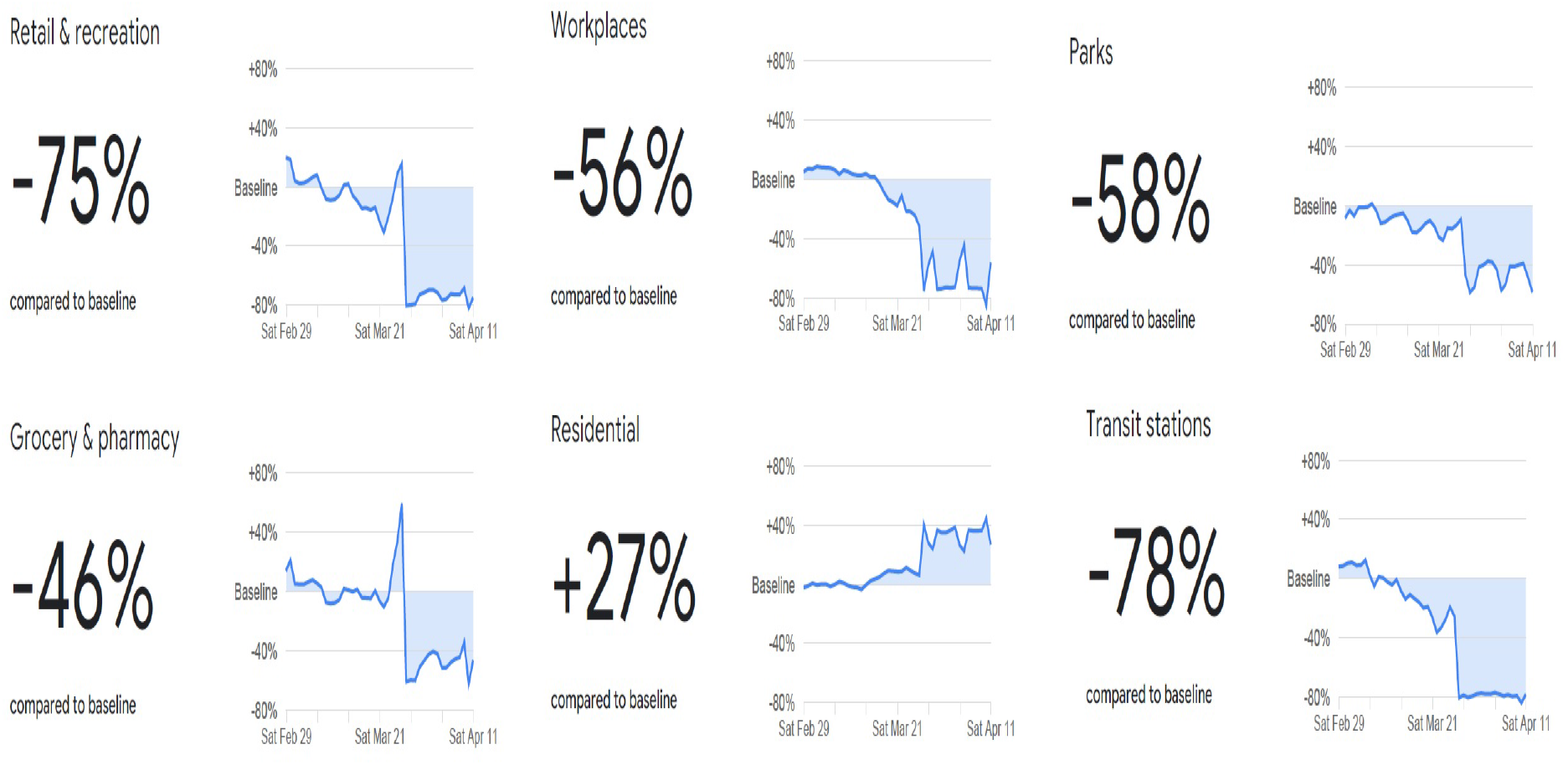
Google mobility data as of 11 April 2020 [9].

**Figure 2:**
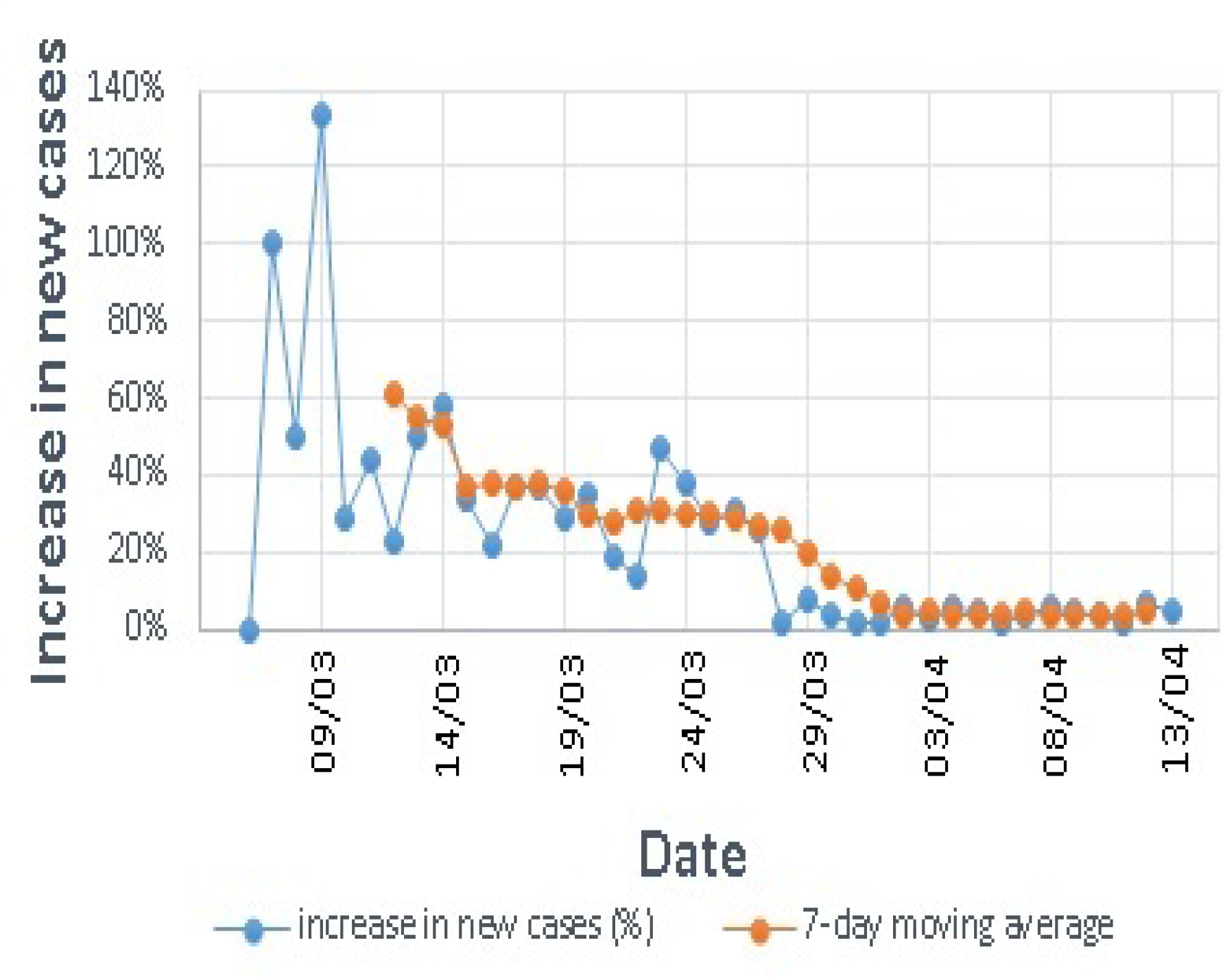
Decrease in new infected cases (%) since the lockdown, together with a 7-day moving average.

### 2.2 Model

We consider an *SEIR* model where the total population *N* (*t*) at any time *t* is divided into the following compartments: The susceptible population *S*(*t*) composed of individuals with no virus in their system but at risk of contracting the infection when they come into contact with infectious individuals, the exposed population *E*(*t*) with individuals who have the virus but not yet able to transmit the virus through sneezing or coughing, the infectious population *I*(*t*) with individuals with either mild or clinical symptoms who can transmit the virus to the healthy individuals resulting in successful infection, and the removed population *R*(*t*) with individuals who can recover either naturally due to robust immune responses or due to supportive treatment available in isolation centres. Thus,

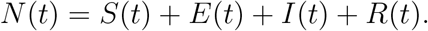

COVID-19, as a lower respiratory infection stays and can be cleared within a few days and so we shall use the day as the unit of time for our model. The vital dynamics for humans are calibrated using the year as a unit of time, hence, converting them into the day unit makes them very small with negligible contribution to the dynamics. We therefore consider a model for COVID-19 with no vital dynamics as a befitting assumption. As stated earlier, the first cases of COVID-19 in South Africa, were due to imported cases, so we assume the flow of individuals into South Africa as both susceptible and exposed, since no infectious cases were picked at the entry point but a few days after entry. We assume a constant immigration rate of Λ. A proportion *p* (0 *< p <* 1) of this immigration rate is assumed to be exposed and 1*− p* is assumed to be susceptible. The recruitment rates of *S*(*t*) and *E*(*t*) are (1 *− p*)Λ and *p*Λ respectively. All the imported exposed cases converted into active cases (*I*(*t*)) within a minimum of 2 days and these active cases were the sources of secondary infections in the country. We assume that the new cases were a result of contacts between the susceptible individuals (*S*(*t*)) with the proportion of individuals who were infectious 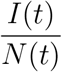 at a rate of successful infection *β*, giving a force of infection 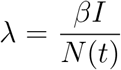 and the number of new infections out of *S*(*t*) and into *E*(*t*) as 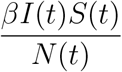. The exposed cases progress to active cases between 2 and 14 days at a constant progression rate *κ* giving the number of individuals moving out of *E*(*t*) and into *I*(*t*) as *κE*(*t*). The infectious individuals are assumed to recover at a constant rate *σ*. The total number of recovered individuals moving out of *I*(*t*) into *R*(*t*) is given by *σI*(*t*). Currently there is no evidence of re-infection after recovery in South Africa and hence, we assume that no recovered individual can become susceptible to the infection.

South Africa implemented a national lockdown on day 21 since the announcement of the first case, where day 1 is March 5 as given in Table 1. The lockdown saw the complete blockage of imported cases and hence, the infection after day 21 was driven by the imported cases which were already in the country until day 21 and the local cases which resulted from these imported cases. After day 21, there were no more recruitment into the susceptible and exposed populations due to immigration as a result of the lock down. This is tantamount to setting both Λ and *p* to zero after day 21. We represent the recruitment rate *S*_*rec*_ by

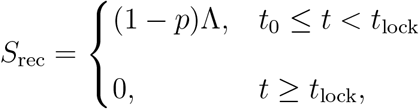

where *t* = *t*_0_ is the time on which the first infectious cases was identified and *t* = *t*_lock_ is the time on which the lockdown was effected. In our model, *t*_0_ = 0 and *t*_lock_ = 21. Similarly, the recruitment rate *E*_rec_ of the exposed population is given by

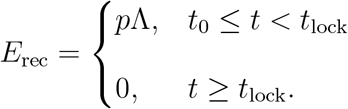

At the inception of the lockdown, the main effective method that was enforced was social distancing between individuals, among other methods. The social distancing aspect reduces the contact rate between the infectious and susceptible individuals. We incorporate social distancing through a constant rate *ρ* (0 *< ρ <* 1), where *ρ* ≃ 0, means that the social distancing is near perfect and *ρ*≃ 1, means there is no social distancing. We assume that *ρ* does not assume its extreme values since the locking down in its inherent nature enforces some social distancing and due to the nature of the settlement patterns in some parts of South African suburbs and the socialisation of family units, perfect social distancing is not attainable. The force of infection is modified as follows:

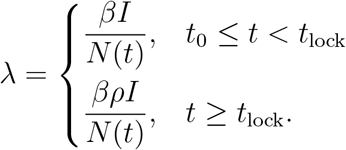

The flow diagram model with the aforementioned assumptions is presented in Figure 3. The system of differential equations capturing the assumptions on COVID-19 for South Africa before and after the lockdown is presented in (1) and given by

**Figure 3:**
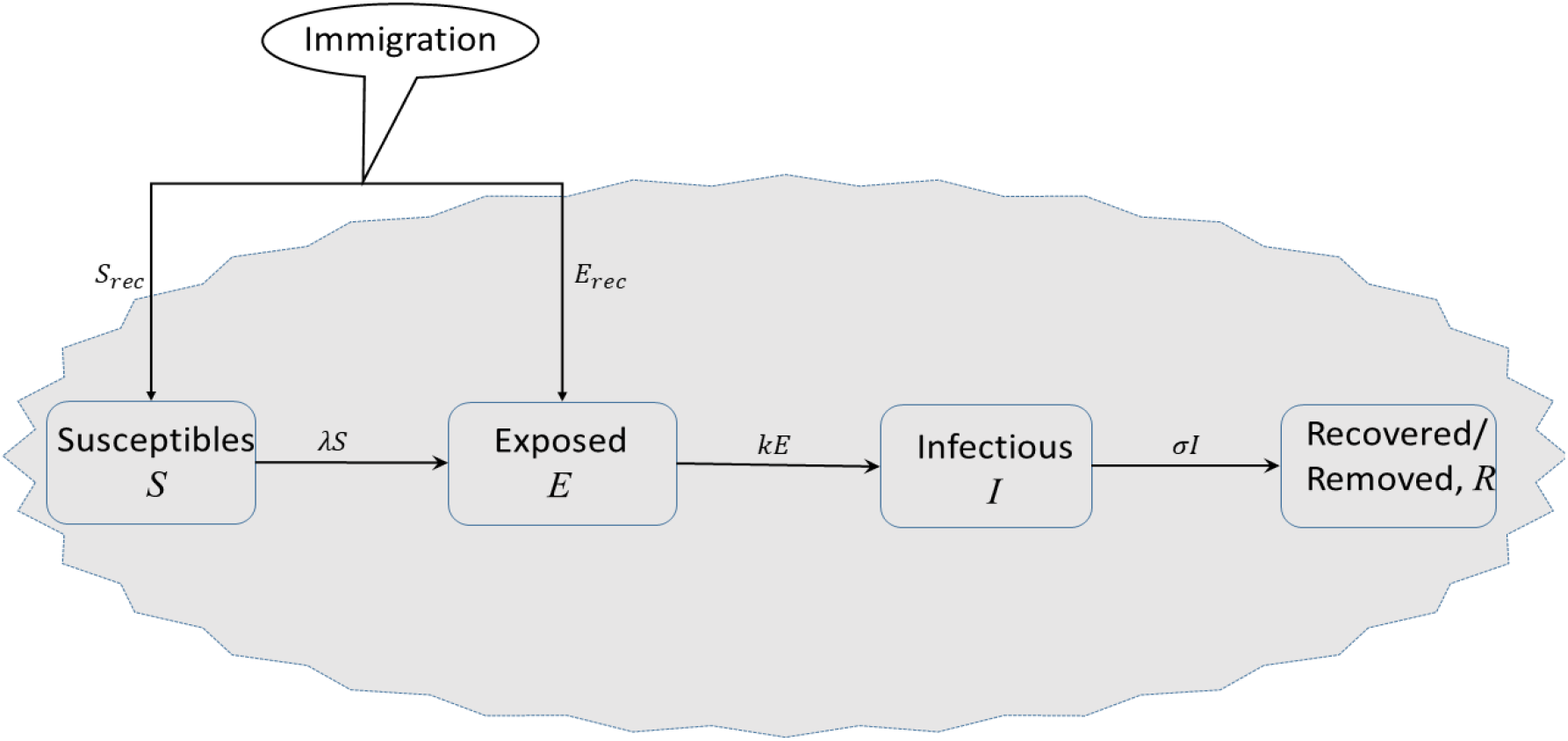
Model diagram for COVID-19 for South Africa with, immigration, lockdown and social distancing.

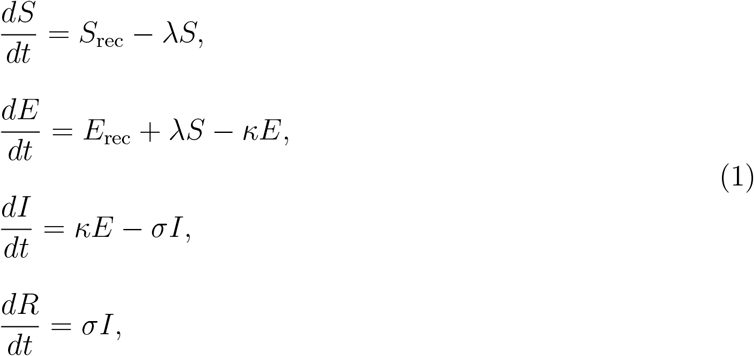

subject to the initial conditions

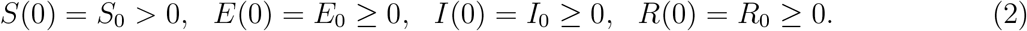

## 3 Results

### 3.1 Data and Parameter Estimation

The epidemic data was obtained from [10] and the information released through the South African government newsroom [11]. According to Statistics South Africa [8], the mid-year population estimates for 2019 showed that the country’s population stood at 58 775 022. This statistic is important in estimating the number of susceptible individuals at the start of the epidemic. It is important to note that at the beginning of the epidemic, the entire country is considered to be naive as the index cases were all as a result of individuals who travelled to places that had already ongoing epidemics such as Asia, Europe and North America.The first case of COVID-19 in South Africa is reported to have travelled to Italy with his wife (as a part of a group of 10) and had returned on the 1^*st*^ of March 2020 [5]. The data on COVID-19 in this study was mainly obtained from the media releases by the South African government [11] and the National Institute of Communicable Diseases [5] from the 5^*th*^ of March to the 13^*th*^ of April 2020. The data included the daily updates on the cumulative number of COVID-19 cases and their distribution per province in some instances. The formulated model is then fitted to the data in two epochs, viz the period before the lockdown and the duration after the lock down. The uniqueness of this model is that it considers a constant daily influx of people into the South African country with a proportion of those coming into the country having been exposed to COVID-19. After the lockdown, the constants that model the movement of people into the country are then set to zero and then the epidemic is assumed to be mainly driven by local transmission dynamics.

We fit the model to the data recorded before the lockdown using the least squares curve fitting method in Matlab. Parameter bounds are set based on recent work on the epidemic in countries such as China, [15,16,19,20], Korea [17,25] and Italy [18]. The reproduction numbers of COVID-19 were determined from earlier studies, see Table 3.

**Table 3:**
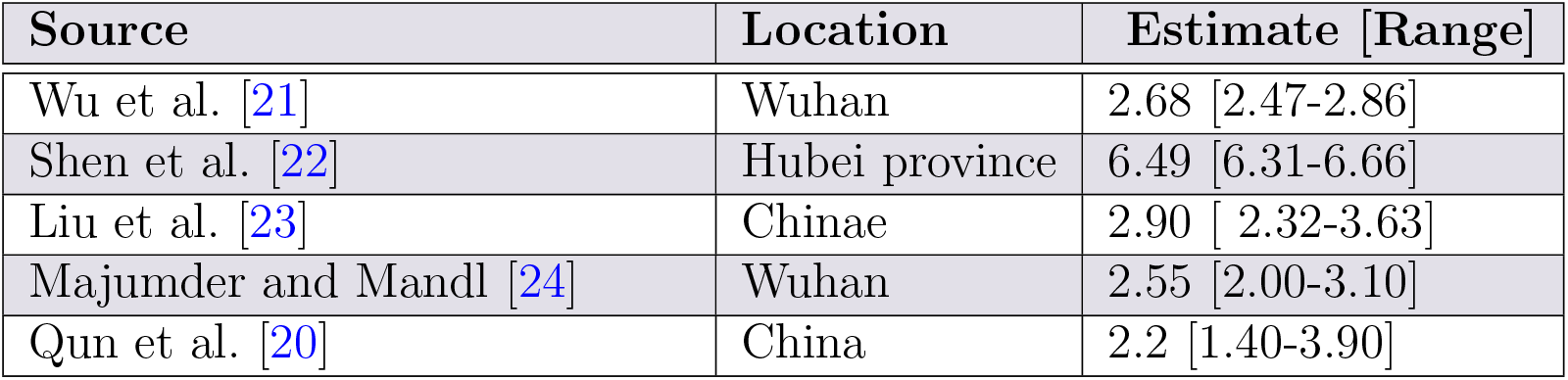
Reproduction numbers derived from various COVID-19 mathematical models for the epidemic in China.

Given the known values of the incubation period, 1*/k*, and the duration of infectivity, 1*/σ*, we estimate the effective contact rate *β*, for the current epidemic in South Africa. The parameter values are given in Table 4.

**Table 4:** Parameters of (COVID-19) *SEIR* model for South Africa and their values.

| Symbol | Parameter description | Value [Range] | Source |
| --- | --- | --- | --- |
| $\beta$ | Effective contact rate | 1.0598 [0.9011-1.400] | Data fit |
| $k$ | Progression rate | 0.5 [0.2-0.6] | [25] |
| $\sigma$ | Recovery rate | 0.4345 [ 0.2-0.5] | [20, 25] |
| $\rho$ | Social distancing parameter | (0,1) | By definition |
| $p$ | Proportion of exposed immigrants | (0,1) | By definition |
| $\Lambda$ | Daily rate of immigration | 11244 [10000-25000] | [26] |

### 3.2 Sensitivity Analysis

The *SEIR* model (1) has some limitations especially on determining the parameter values. The various processes driving the model dynamics have a number of complexities and hence, the model parameters are best measured approximately. The parameters have substantial variations depending on geographical region, demographic factors, and several other factors. In addition, some of the parameters may just be stochastic in nature. The model thus, has inherent epistemic uncertainty which result from lack of knowledge about the value of parameters which are then assumed to be constant throughout the model analysis [27]. To adapt the model simulations to the South African scenario, we first used parameter values from other regions, fitted the model to the daily cumulative cases and obtained the optimal values of the parameters. To cater for the inherent complexities around these optimal values, we use the Latin Hypercube Sampling (LHS) technique for uncertainty quantification and sensitivity analysis. This is a reliable and efficient technique for detection of such epistemic uncertainties allowing unbiased estimates of the average model output with fewer samples than other sampling methods to achieve the same accuracy. To measure the strength of the relationship between each input variable and each output variable, the LHS technique is combined with the partial rank correlation coefficients (PRCCs) [28]. We use Figures 4 and 5 to investigate the contribution of different parameters before and after the lock-down. Their contribution is measured against the number of cumulative cases in South Africa. The range of values used are given in Table 3. Figure 4 shows that the immigration parameters *p* and Λ are strongly positively correlated to the number of cumulative cases before the lockdown. This means that the increase in the proportion of exposed immigrants and the rate of immigration are associated with the increase in cumulative cases. Figure 5 shows that the infection parameter *β* and the social distancing parameter *ρ* are strongly positively correlated to the cumulative cases whilst the infection progression and recovery parameters *κ* and *σ* are strongly negatively correlated to the cumulative cases after lockdown. The increase in the infection rate and the increase in the social distancing parameter (i.e. decreasing social distancing level) is associated with the increase in cumulative parameters whilst the increase in the progression and recovery rates are associated with the decrease in cumulative cases.

**Figure 4:**
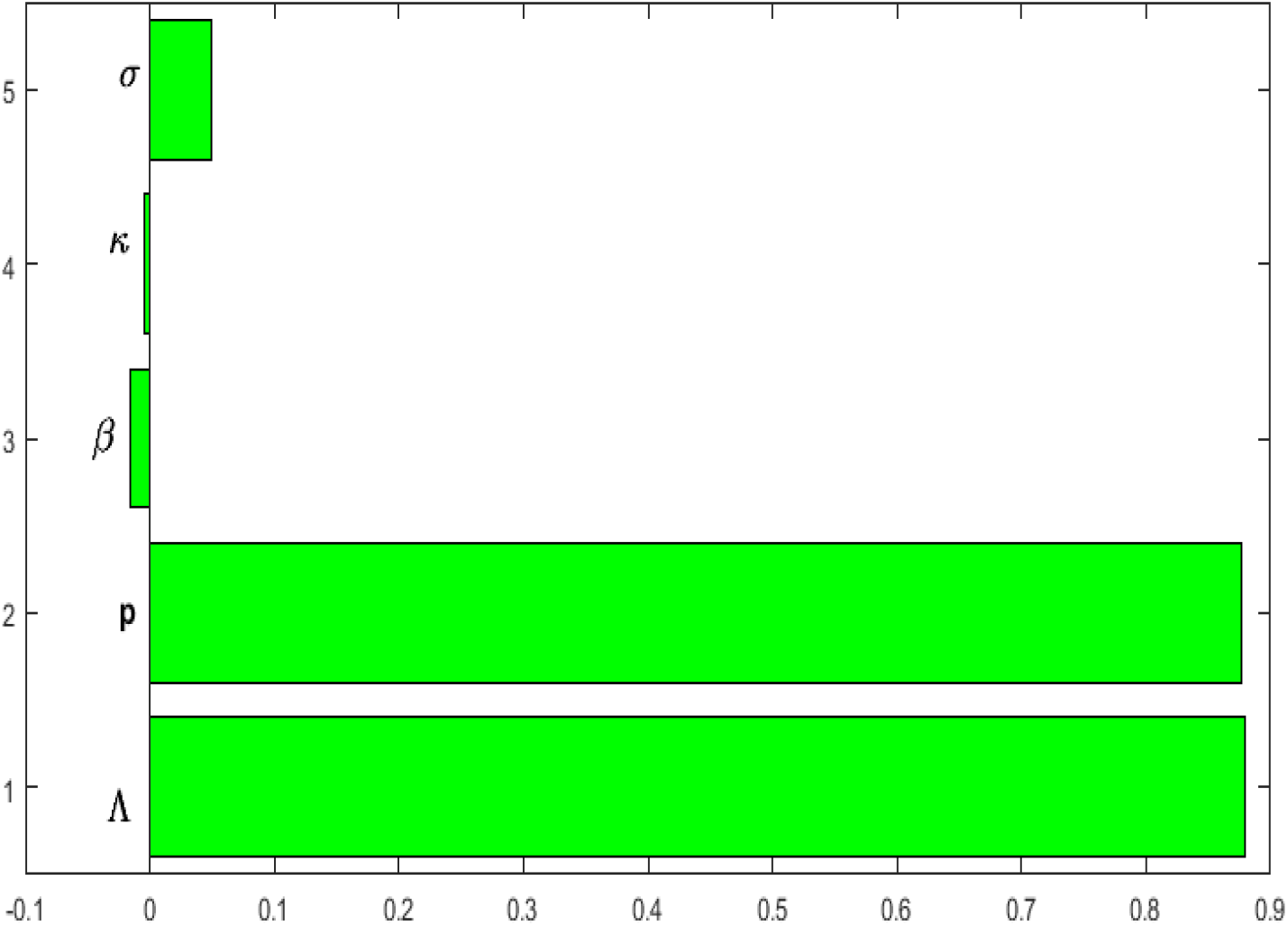
Influence of parameter values on the cumulative cases before the lockdown.

**Figure 5:**
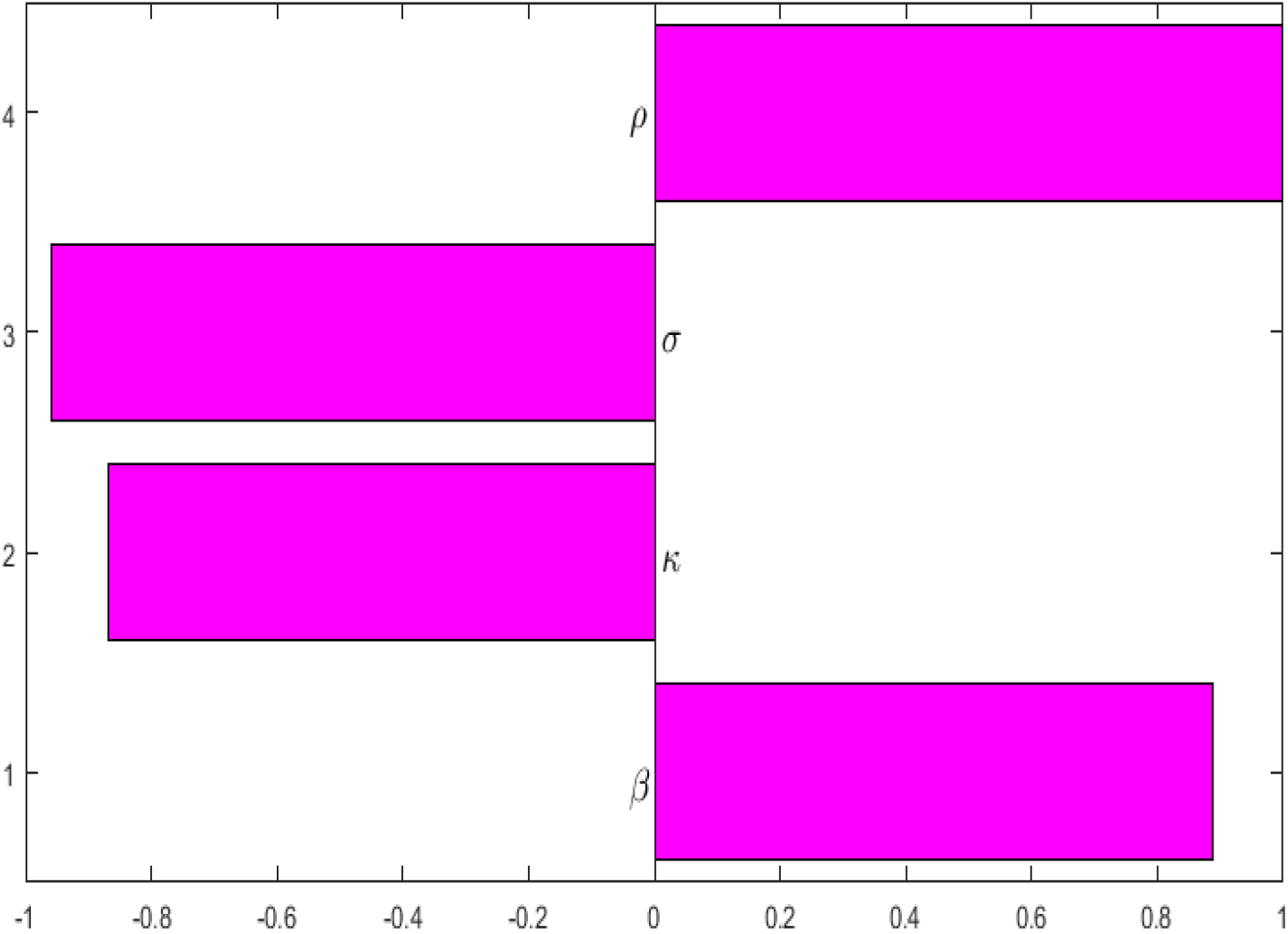
Influence of parameter values on the cumulative cases after the lockdown

### 3.3 Model Fitting and Simulations

In this section we fit the designed model to the cumulative cases of South Africa before the lockdown (see Figure 6) in the absence of any intervention to obtain the optimal fit and optimal model parameter values for the South African dynamics. We present the model simulations with these optimal parameter values to predict the likelihood of occurrence of the peak number of cases had the lock-down not been effected at day 21 (Figures 7 and 8). We also fit the model to the cumulative cases using when lockdown and social distancing (Figure 9) were effected to obtain the optimal parameter values for the fit. We use the resulting fit to make possible predictions of the potential impact of different levels of compliance to social distancing with the lockdown in place (Figure 10).

**Figure 6:**
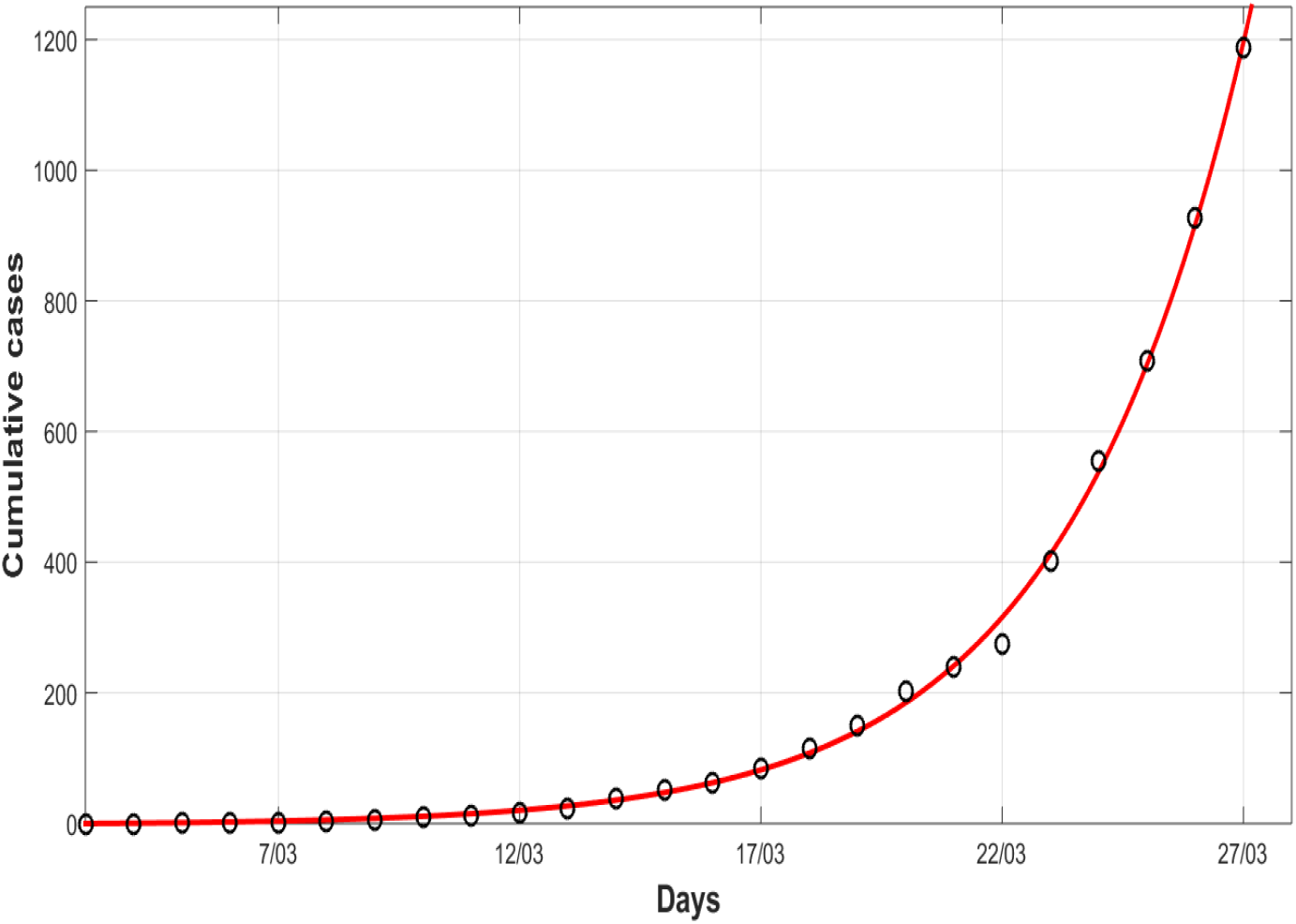
Model fit to data for COVID 19 before the lockdown. Optimal parameter values: Λ = 11244, *p* = 1.7598 *×* 10^*−*5^, *β* = 1.1411, *κ* = 0.655, *σ* = 0.4482, *ρ* = 1.

**Figure 7:**
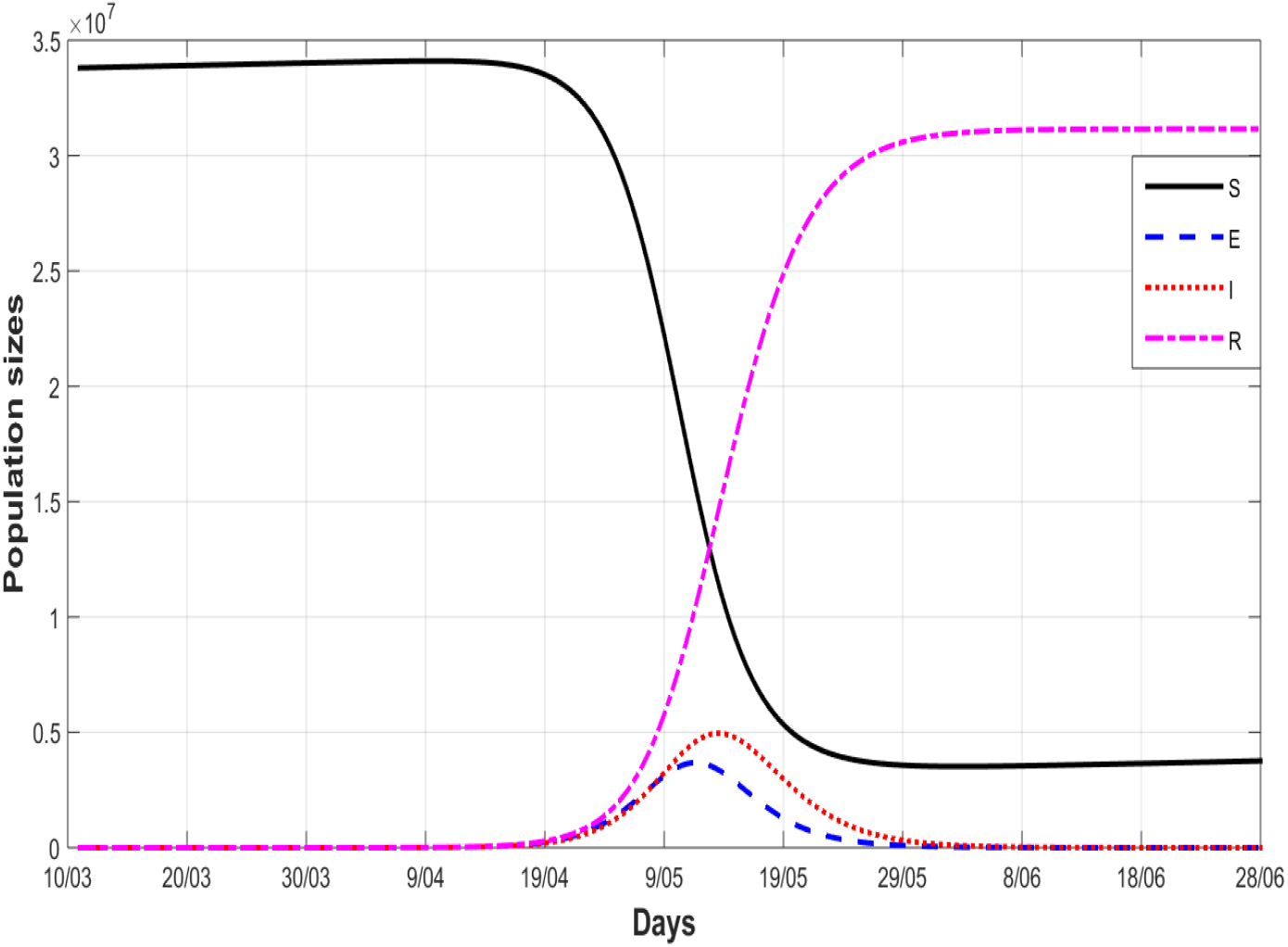
A simulation model for the *SEIR* model for all the populations in the absence of any intervention, i.e *ρ* = 1. Optimal parameter values: Λ = 11244, *p* = 1.7598 *×*10^*−*5^, *β* = 1.1411, *κ* = 0.655, *σ* = 0.4482.

**Figure 8:**
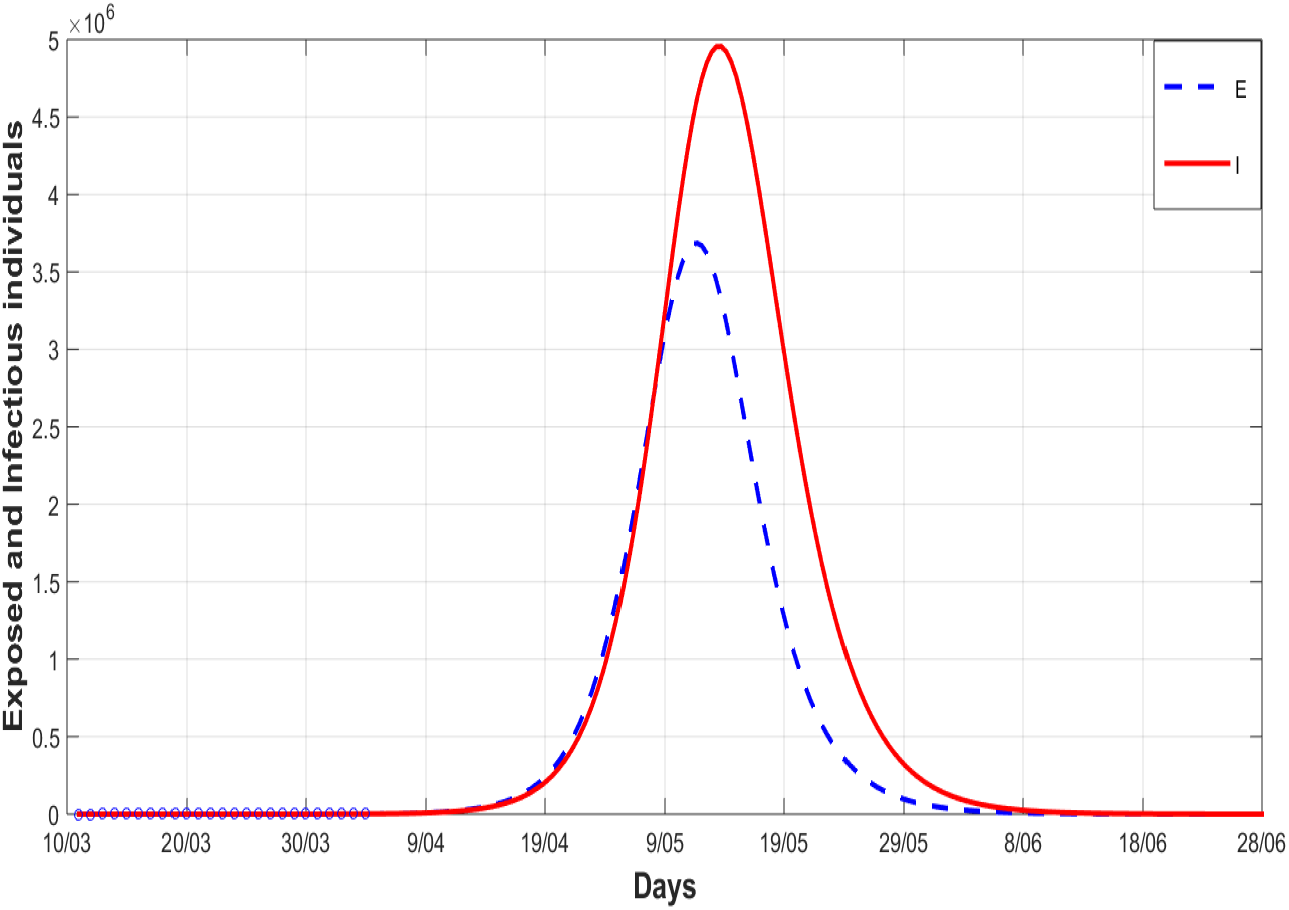
A simulation model for the *SEIR* model for exposed and infected in the absence of any intervention i.e *ρ* = 1. Optimal parameter values: Λ = 11244, *p* = 1.7598 *×* 10^*−*5^, *β* = 1.1411, *κ* = 0.655, *σ* = 0.4482.

**Figure 9:**
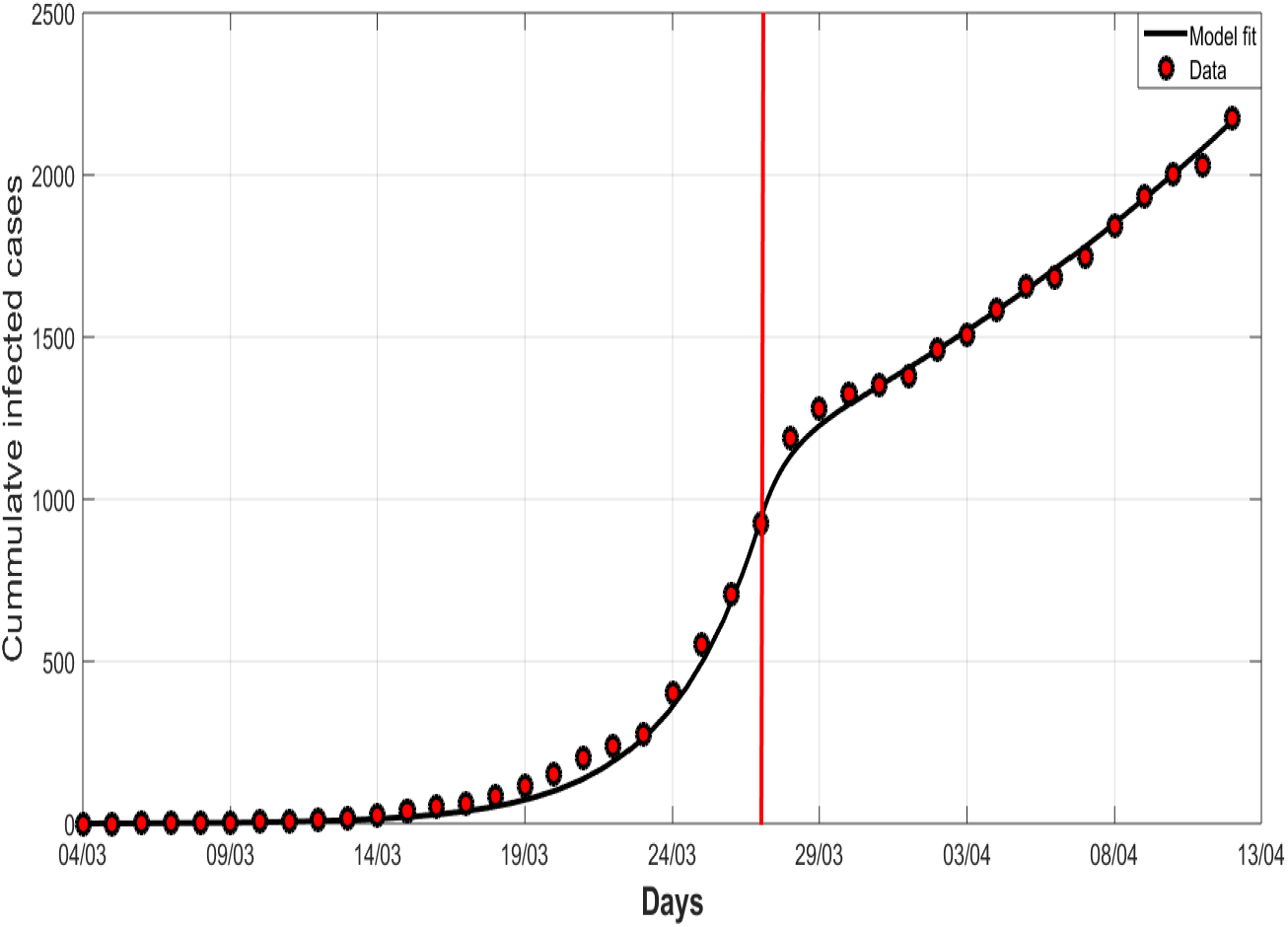
The graph shows COVID-19 model fitting for the cumulative infected cases for South Africa before and after lockdown. The red dots mark are the cumulative reported number of COVID-19 cases before and after lockdown. The red vertical line represents the start of the national lockdown. The continuous curve represent the model fit. Optimal parameter values: Λ = 11244, *p* = 1.7598*×* 10^*−*5^, *β* = 1.1411, *κ* = 0.655, *σ* = 0.4482, *ρ* = 0.453. Note that *ρ* = 0.453 represents about 55% compliance in social distancing.

**Figure 10:**
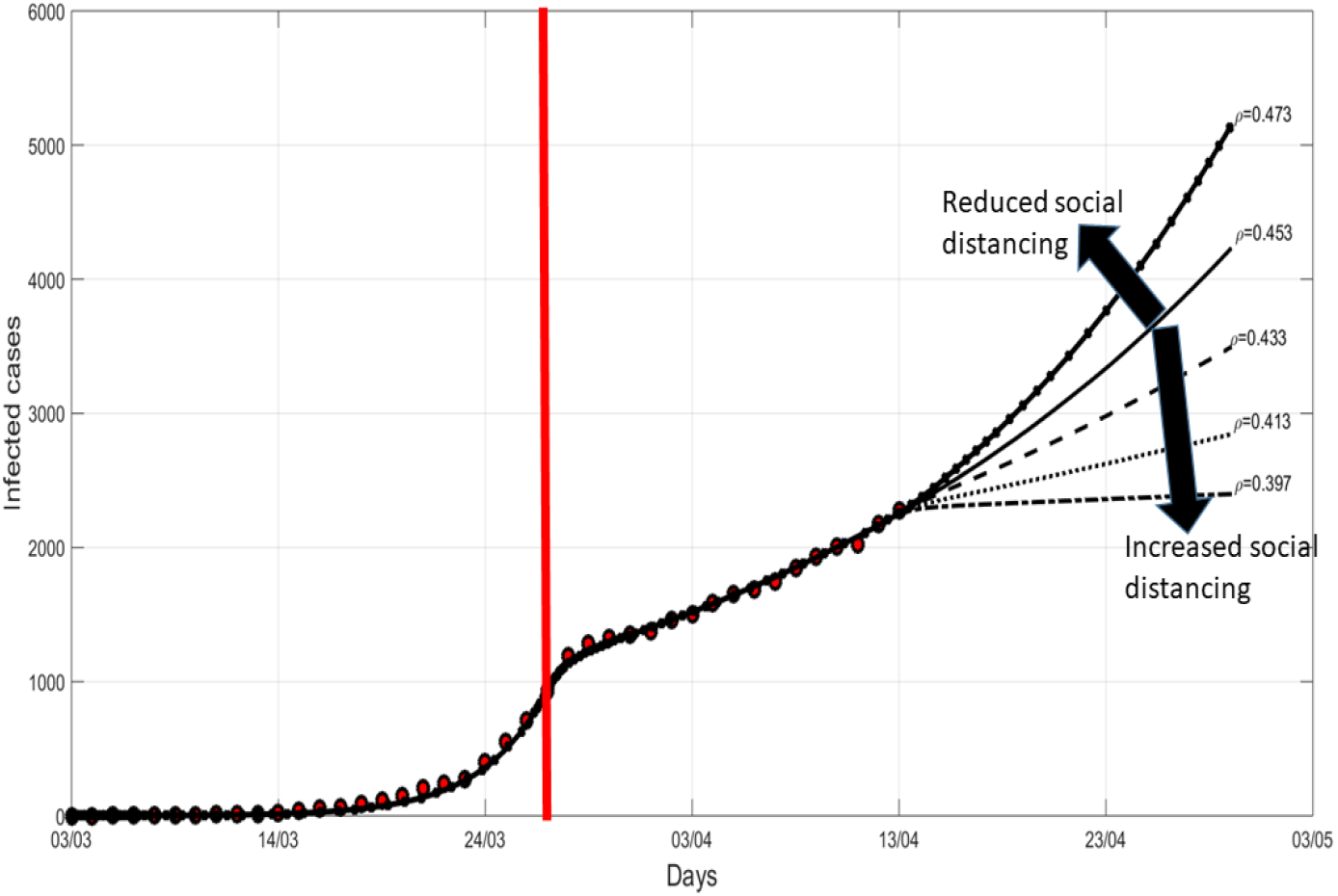
The graph shows COVID-19 model fitting for the cumulative infected cases for various scenarios. The trajectories for different levels of social distancing are shown by the different line types. The red dots mark are the cumulative reported number of COVID 19 cases in South Africa before the lockdown and two weeks after the lock-down. The red vertical line denotes the time when a lockdown was implemented by the government with a delay of one data point as the reported cases represent posterior data. Optimal parameter values are: Λ = 11244, *p* = 1.7598 *×* 10^*−*5^, *β* = 1.1411, *κ* = 0.655, *σ* = 0.4482, *ρ* = 0.453.

Figure 6 shows that the model (1) fits well to the cumulative cases of South Africa and the optimal parameter values are the current model values that could be used for prediction in the absence of intervention. Using the optimal parameter values, Figure 7 shows that the peak of cumulative cases would have been reached in mid-May before the of cases begin to decline. (Figures 8 is an extract of Figure 7 showing the quantum of exposed and infectious populations in the absence of any intervention. The figure shows that there would be more infectious cases at the peak than the exposed cases by the time the peak is reached.

Figure 9 shows that the model (1) fits well to the cumulative cases of South Africa up to the 13^*th*^ of April 2020 in the presence of intervention and the optimal parameter values are the current model values that could be used for prediction in the presence of intervention. We note that the optimal fit for social distancing after lock-down is about 55%(*ρ* = 0.45). Figure 10 shows COVID-19 model fitting and predictions for the cumulative infected cases for various levels of social distancing. The simulations show that if the optimal level of social distancing is maintained at 55%, then the number of cumulative cases will continue to grow exponentially. Reducing the level of social distancing from 55% to 53% (*ρ* = 0.47) would increase the number of cumulative cases by about 23% at the end of lock-down on 30 April 2020. Increasing the level of social distancing from 55% to 57% (*ρ* = 0.43), 59% (*ρ* = 0.41) and 61% (*ρ* = 0.39) would avert the cumulative cases by about 18%, 32% and 53% respectively at the end of lockdown on 30 April 2020. Our model fitting and predictions suggest that the current level of social distancing as at 13 April under lockdown for South Africa is not good enough to flatten the curve and any relaxation from this will lead to a spike in the cumulative cases. The predictions also suggest that efforts to realise the flattening of the curve should be sustained at least at 60%(*ρ* = 0.4) level of social distancing by the time the lockdown ends on 30 April 2020.

## 4 Conclusion

The demand for assessment of methods of control for COVID-19 in South Africa is overwhelming. Understanding the impact of these control measures requires knowledge and expertise drawn from various scientific disciplines including mathematical modelling. The role of mathematical models’ insights formed the national response to the pandemic. The idea of social distancing in order to flatten the epidemic growth curve, thus allowing time to prepare for the worst case scenarios, was drawn from mathematical models for the COVID-19 epidemic. This was critical in preventing as many deaths as possible.

We presented a deterministic model to assess the impact of social distancing on the transmission dynamics of COVID-19 in South Africa taking into account the initial imported cases before the lockdown in the early phase of infection. The model fitted well to the South African cumulative cases data leading to an appropriate estimation of the optimal model-based parameter estimates for South Africa. Insights gained from the current study contributes impacts the current levels of intervention using social distancing in South Africa. The results suggest that individuals migrating into South Africa played an important role in driving the infection in South Africa and the prevention of infections happening elsewhere on the globe needs special attention due to improved mobility of humans. Particular attention need to be focused on this group of people beyond the lockdown to ensure that only negative cases are allowed into the country.

The sensitivity results show the shift in the drivers of infection before and after implementation of interventions. This may imply that any intervention leads to emerging challenges due to shifting in the drivers and hence proper planning is needed to scale up efforts to avert any emerging issues. Since all the parameters after lock down have strong correlation to the number of cumulative cases, holistic strategies are required to reduce the processes increasing the number of cases simultaneously with those decreasing the number of cumulative cases.

The current levels of social distancing were predicted to be inadequate especially during this infant stage of infection with exponential growth. There is need for more aggressive and robust multi-control approaches that target reduction of infection rate, increase of social distancing levels, rapid detection of exposed cases and increasing the recovery of active cases need to be implemented simultaneously and optimised. The shifting in drivers of infection after implementation of lockdown imposes threat to the preparedness of the South African system even in the aftermath of the lock-down. The use multidisciplinary collaboration and predictive tools is needed to forewarn the South African systems of such shifts and enhance emergency preparedness responses.

The model presented here is not without shortcomings. The dynamics of COVID-19 combined with the economic and social dynamics of South Africa present a complex scenario from a modelling perspective. On the social dynamics, social distancing in many of the informal settlements has been difficult to enforce due to the high density of people per unit area. In addition, the economic dynamics in many of the informal settlements is influenced by informal trading, thus creating difficulties and hardships to people during a lockdown. There were relaxations of the social distancing measures after the lockdown to mitigate the economic and social challenges of the general population. These relaxations had the potential to dilute the intended effects of social distancing. The model presented in this paper was simple and did not capture these complexities. Despite the shortcomings, the results present some interesting results in which even after the simulations, the data that was released on the epidemic still fell within the predictions of the model.

## Data Availability

The data is secondary data available in the public domain.

https://github.com/dsfsi/covid19za

## Author Contributions

FN, WC and FC were responsible initial manuscript preparation. FN and FC were involved in the numerical simulations and their write up. MVV assisted with the conceptualisation of the paper and preparation of the manuscript. All authors were involved in writing the manuscript.

## Data Availability

The data are sourced from public and official government press release [10, 11].

## Acknowledgements

The authors would like to thank the Faculty of Science in the University of Johannesburg for material support towards the inception and completion of this project.

